# Expert-Level Detection of Referable Glaucoma from Fundus Photographs in a Safety Net Population: The AI and Teleophthalmology in Los Angeles Initiative

**DOI:** 10.1101/2024.08.25.24312563

**Authors:** Van Nguyen, Sreenidhi Iyengar, Haroon Rasheed, Galo Apolo, Zhiwei Li, Aniket Kumar, Hong Nguyen, Austin Bohner, Rahul Dhodapkar, Jiun Do, Andrew Duong, Jeffrey Gluckstein, Kendra Hong, Lucas Humayun, Alanna James, Junhui Lee, Kent Nguyen, Brandon Wong, Jose-Luis Ambite, Carl Kesselman, Lauren Daskivich, Michael Pazzani, Benjamin Y. Xu

## Abstract

**Purpose:** To develop and test a deep learning (DL) algorithm for detecting referable glaucoma in the Los Angeles County (LAC) Department of Health Services (DHS) teleretinal screening program.

**Methods:** Fundus photographs and patient-level labels of referable glaucoma (defined as cup-to-disc ratio [CDR] ≥ 0.6) provided by 21 trained optometrist graders were obtained from the LAC DHS teleretinal screening program. A DL algorithm based on the VGG-19 architecture was trained using patient-level labels generalized to images from both eyes. Area under the receiver operating curve (AUC), sensitivity, and specificity were calculated to assess algorithm performance using an independent test set that was also graded by 13 clinicians with one to 15 years of experience. Algorithm performance was tested using reference labels provided by either LAC DHS optometrists or an expert panel of 3 glaucoma specialists.

**Results:** 12,098 images from 5,616 patients (2,086 referable glaucoma, 3,530 non-glaucoma) were used to train the DL algorithm. In this dataset, mean age was 56.8 ± 10.5 years with 54.8% females and 68.2% Latinos, 8.9% Blacks, 2.7% Caucasians, and 6.0% Asians. 1,000 images from 500 patients (250 referable glaucoma, 250 non-glaucoma) with similar demographics (p ≥ 0.57) were used to test the DL algorithm. Algorithm performance matched or exceeded that of all independent clinician graders in detecting patient-level referable glaucoma based on LAC DHS optometrist (AUC = 0.92) or expert panel (AUC = 0.93) reference labels. Clinician grader sensitivity (range: 0.33-0.99) and specificity (range: 0.68-0.98) ranged widely and did not correlate with years of experience (p ≥ 0.49). Algorithm performance (AUC = 0.93) also matched or exceeded the sensitivity (range: 0.78-1.00) and specificity (range: 0.32-0.87) of 6 LAC DHS optometrists in the subsets of the test dataset they graded based on expert panel reference labels.

**Conclusions:** A DL algorithm for detecting referable glaucoma developed using patient-level data provided by trained LAC DHS optometrists approximates or exceeds performance by ophthalmologists and optometrists, who exhibit variable sensitivity and specificity unrelated to experience level. Implementation of this algorithm in screening workflows could help reallocate eye care resources and provide more reproducible and timely glaucoma care.

## Introduction

Glaucoma is the leading cause of irreversible blindness worldwide with prevalence growing from 64.3 million in 2013 to 111.8 million in 2040.^1,2^ In the United States, glaucoma is projected to affect 7.3 million people by 2050 with the majority being racial minorities.^2^ The rising burden of glaucoma in the United States is exacerbated by a critical shortage of eye care providers; the total supply of ophthalmologists is projected to decrease by 12% while demand for eye care services is projected to increase by 24% by 2035.^3^ Underserved racial minorities and individuals living in non-metro areas who already experience difficulty accessing care will likely be disproportionately affected, thereby exacerbating ongoing disparities in glaucoam care.^7^ For example, Blacks and Hispanics in the US have a significantly higher risk of glaucoma-related blindness and need for glaucoma surgery compared to non-Hispanic Whites.^4–7^ Therefore, there is an urgent need to develop and implement novel interventions that address the impending eye care crisis by ensuring timely and equitable detection of at-risk individuals.

The Los Angeles County (LAC) Department of Health Services (DHS), the second largest municipal health system in the United States, has operated a teleretinal screening program for newly diagnosed diabetics since 2013.^19^ While the program primarily focuses on detecting diabetic retinopathy, it also screens for other ocular conditions, including cataracts and referable glaucoma. The referable glaucoma component of the program has been effective; between 2016 to 2018, 817 patients were referred for glaucoma evaluations, 534 (65.4%) patients successfully completed in-person evaluations, and 131 (24.5%) patients were diagnosed with glaucoma by LAC DHS clinicians.^8^ Despite its success, the program is hindered by key workflow limitations. Reliance on trained optometrists to manually grade fundus photographs contributes to referral delays and takes time away from direct patient care. Manual grading by over 20 LAC DHS optometrists also potentially introduces inter-grader variability in disease detection.^23^ Therefore, it is critical to consider alternative approaches for standardizing and streamlining referrals to ensure reproducibility and equity of care.

Artificial intelligence (AI), specifically deep learning (DL), is an emerging technology in healthcare that could enhance the reproducibility and efficiency of glaucoma screening, thereby enabling earlier detection and intervention. In this study, we develop a DL algorithm for detecting referable glaucoma from optic nerve photos of patients in LAC DHS teleretinal screening program. We also perform a rigorous validation of the algorithm by comparing its performance to a panel of 13 eye care providers, including 4 fellowship-trained glaucoma specialists. This type of algorithm, once rigorously validated against standard-of-care human grading, could be implemented to address the critical need for reproducible and scalable solutions in glaucoma screening, especially among vulnerable, safety net populations.

## Methods

This study was approved by the University of Southern California Institutional Review Board. The study adhered to the tenets of the Declaration of Helsinki and complied with the Health Insurance Portability and Accountability Act.

### Data Source

The LAC DHS health system administers a remote teleretinal screening program across 17 hospital- and community-based sites across Southern California.^24^ The program serves around 1,750 newly diagnosed diabetics per month. LAC DHS patients participating in the program receive dilation and fundus photography by trained technicians using the Topcon NW400 and NW8 (Topcon Corporation, Tokyo, Japan) and Canon CR-2 AF Digital (Canon U.S.A. Inc, Huntington, NY) cameras. These photographs are evaluated primarily for diabetic retinopathy and secondarily for referable glaucoma, defined as cup-to-disc ratio (CDR) ≥ 0.6, by 21 trained LAC DHS optometrists. Disease diagnoses, including for referable glaucoma, are recorded on the patient level. All patients 18 years of age or older with at least one fundus photograph taken between January 4, 2016 to December 2, 2022 were eligible for analysis.

A segmentation-free approach to detecting referable glaucoma was selected given: 1) generally superior diagnostic performance compared to segmentation-reliant approaches; and 2) lack of access to CDR and segmentation data in the LAC DHS dataset.^9^ Fundus photos centered on the optic nerve from all patients diagnosed with referable glaucoma and a comparable number of patients diagnosed as non-glaucoma were retrieved for purposes of AI algorithm development. All photos underwent went manual review. Photos of low quality (e.g. blurry, underexposed or overexposed, or media opacities partially obscuring the optic nerve) were included to ensure generalizability of algorithms to real-world screening environments. However, photos were excluded if they could not be graded for glaucoma (e.g. media opacities totally obscuring the optic nerve, so out of focus that the optic nerve could not be delineated, or if the optic nerve was not in the field of view).

Fundus photos were cropped and centered around the optic nerve head for analysis in a two-step process that was programmed in Python. First, the program cropped each raw fundus image to the image region by removing any black or extraneous regions. Then, the program scanned the image using a sliding window approach that attempted to match the cropped image to the pattern of an optic disc. Once a potential match was found, that section of the image was saved as the final cropped image. If the program failed to locate or confirm an optic disc after multiple attempts, the entire uncropped image was saved. All images were manually reviewed to ensure cropping and centration were effective. Images where the optic disc was present but difficult to visualize due to occlusion or exposure issues were retained in the dataset to represent real-world scenarios. Images without an optic disc were excluded. Images were resized to 224 by 224 pixels to reduce hardware demands during training. Images were preprocessed by normalizing RGB channels and augmented through random rotation, translation, and perturbations to balance and contrast.

### Algorithm Development and Validation

The LAC DHS dataset was divided into development (80%) and test (20%) datasets. The development dataset was further split into training (75%) and validation (25%) datasets. Some patients with multiple teleretinal screening visits were represented multiple times in the training and validation datasets, although reference labels by LAC DHS optometrists were unique for each visit. The test dataset was used to derive a sample of 1000 test images from 500 patients with no overlap of patients with the training or validation datasets.

Patient-level labels of referable glaucoma were provided by one of 21 trained LAC DHS optometrists after analyzing photos of both eyes. These patient-level labels were generalized to photos from both eyes to train DL algorithms for detecting referable glaucoma on the eye level. A convolutional neural network (CNN) was developed based on the VGG-19 architecture using the training and validation datasets labeled in this manner. The VGG-19 architecture was chosen due to its efficiency with image-based data while providing similar performance to other architectures, including InceptionV3, MobileNetV3, EfficientNetV2, and ResNet50V2. The average pooling layer was replaced by an adaptive pooling layer where bin size is proportional to input image size, enabling the CNN to be applied to input images of arbitrary sizes. Softmax-regression was used to calculate the multinomial probability of the three classes with a cross-entropy loss used during training. All layers of the CNN were fine-tuned using backpropagation; optimization was performed using stochastic gradient descent with warm restarts. The algorithm was then fine-tuned using transfer learning from labels provided by a glaucoma specialist.

The DL algorithm was tested using the 1000-image test dataset, which was also graded by 13 clinicians (1 optometrist, 7 general ophthalmologists, 1 neuro-ophthalmologist, and 4 glaucoma specialists) with between one to 15 years of clinical experience. Prior to grading, each of these clinicians was provided with a standardization dataset comprised of 20 images per CDR between 0.2 to 0.9 in 0.1-unit increments. As one objective of the study was to assess the effect of provider experience, the size of the sample dataset was limited to avoid strongly biasing providers with less experience.

Three sets of reference labels of the independent test set were used to assess algorithm performance. The LAC DHS optometrist reference labels were provided on the patient level by 21 trained LAC DHS optometrists who originally graded the photos in the test dataset. Expert panel reference labels were provided by 3 of the fellowship-trained glaucoma specialists (V.N., B.W., B.Y.X.) among the 13 study graders, with their majority diagnosis (at least 2 of 3) determining the glaucoma status for each individual photo. Expert panel reference labels were provided on the eye level and combined to generate patient-level labels; a patient was positive for referable glaucoma if at least one eye was labeled as such.

### Data Analysis

Demographic characteristics between the training/validation and test sets were compared using a 2-tailed student t-test or a chi-square test. The study cohort was stratified by glaucoma status based on LAC DHS optometrist labels to compare demographic and clinical characteristics. Continuous measures were summarized by means and standard deviations and categorical measures were summarized by proportions and percentages. Area under the receiver operating characteristic curve (AUC), sensitivity, and specificity were calculated to assess algorithm performance compared to the sensitivity and specificity of individual clinician graders using all three sets of reference labels. A sub-analysis comparing LAC DHS optometrist and algorithm performance was performed for the LAC DHS six optometrists who graded the largest proportion of the independent test set using the expert panel reference labels. Linear regression was performed to assess the association between grader sensitivity or specificity and years of clinical experience. Statistical tests were considered statistically significant when the P value < 0.05. Statistical analyses were performed using Python’s SciPy statistics library.

## Results

A total of 13,098 images were retrieved, and 12,998 images were included in the analysis after excluding 100 cropped images (0.76%) without visible optic nerves. The training dataset had 8,996 images from 4,212 patients, the validation dataset had 3,002 images from 1,404 patients, and the test dataset had 1,000 images from 500 patients. The 5,616 patients (2,086 referable glaucoma, 3,530 non-glaucoma) in the training and validation datasets had mean age of 56.8 ± 10.5 years, and there were 54.8% (N = 3091) females, 68.2% (N = 3826) Latinos, 8.9% (N = 501) Blacks, 2.7% (N = 153) Caucasians, 6.0% (N = 338) Asians, and 14.2% (N = 798) Other or Not Specified race (Table 1). The 500 patients (250 referable glaucoma, 250 non-glaucoma) in the test dataset had mean age of 57.3 ± 10.3 years, and were 52.4% (N = 262) females, 69.2% (N = 346) Latino, 8.6% (N = 43) Black, 2.6% (N = 13) Caucasian, 5.2% (N = 26) Asian, and 15.0% (N = 75) Other or Not Specified race (Figure 1). There was no difference in age (p = 0.295), race (p = 0.781), or sex (p = 0.569) between patients in the training/validation and test datasets (Table 1).

**Figure 1.**
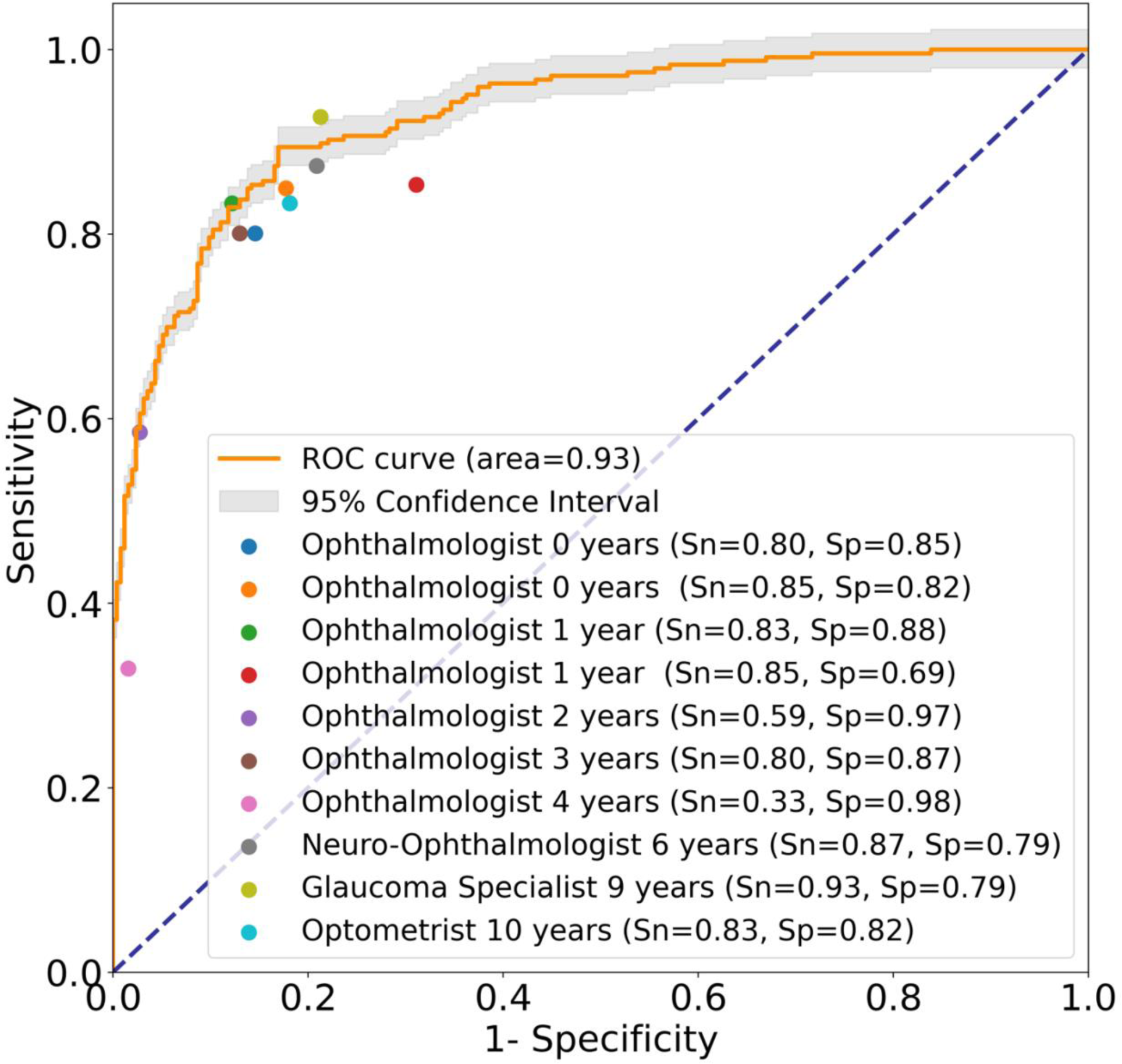
Patient-level algorithm and independent clinician performance (with years of experience) when using patient-level expert panel reference labels. Sn = Sensitivity; Sp = Specificity.

**Table 1.** Baseline demographics stratified by training/validation or test dataset.

| Parameter | Training/Validation | Test | P-value |
| --- | --- | --- | --- |
| Age | 56.8 ± 10.5 | 57.3 ± 10.3 | 0.30 |
| Sex |  |  | 0.57 |
| Female | 55.0% (N = 3091) | 52.4% (N = 262) |  |
| Male | 42.8% (N = 2401) | 44.8% (N = 224) |  |
| Race |  |  | 0.78 |
| Latinos | 68.1% (N = 3826) | 69.2% (N = 346) |  |
| Blacks | 8.9% (N = 501) | 8.6% (N = 43) |  |
| Caucasians | 2.7% (N = 153) | 2.6% (N = 13) |  |
| Asians | 6.0% (N = 338) | 5.2% (N = 26) |  |
| Other or Not Specified | 14.2% (N = 798) | 15.0% (N = 75) |  |
| Glaucoma status |  |  | < 0.001 |
| Referable | 37.1% (N = 2086) | 50.0% (N = 250) |  |
| Non-Referable | 62.9% (N = 3530) | 50.0% (N = 250) |  |

Statistical significance tested by 2-tailed student t-test or Chi-squared test.

Algorithm performance for detecting referable glaucoma on the patient level based on expert panel labels of the test dataset had an AUC of 0.93 (95% CI, 0.91-0.95), with a sensitivity of 0.89 and specificity of 0.83. Individual graders had a sensitivity ranging from 0.33 to 0.99 and a specificity ranging from 0.68 to 0.98, including a sensitivity of 0.98 and specificity of 0.79 by a 4^th^ glaucoma specialist (Figure 1). There was no association between years of clinical experience and grader sensitivity (p = 0.491) or specificity (p = 0.559) (Figure 2).

**Figure 2.**
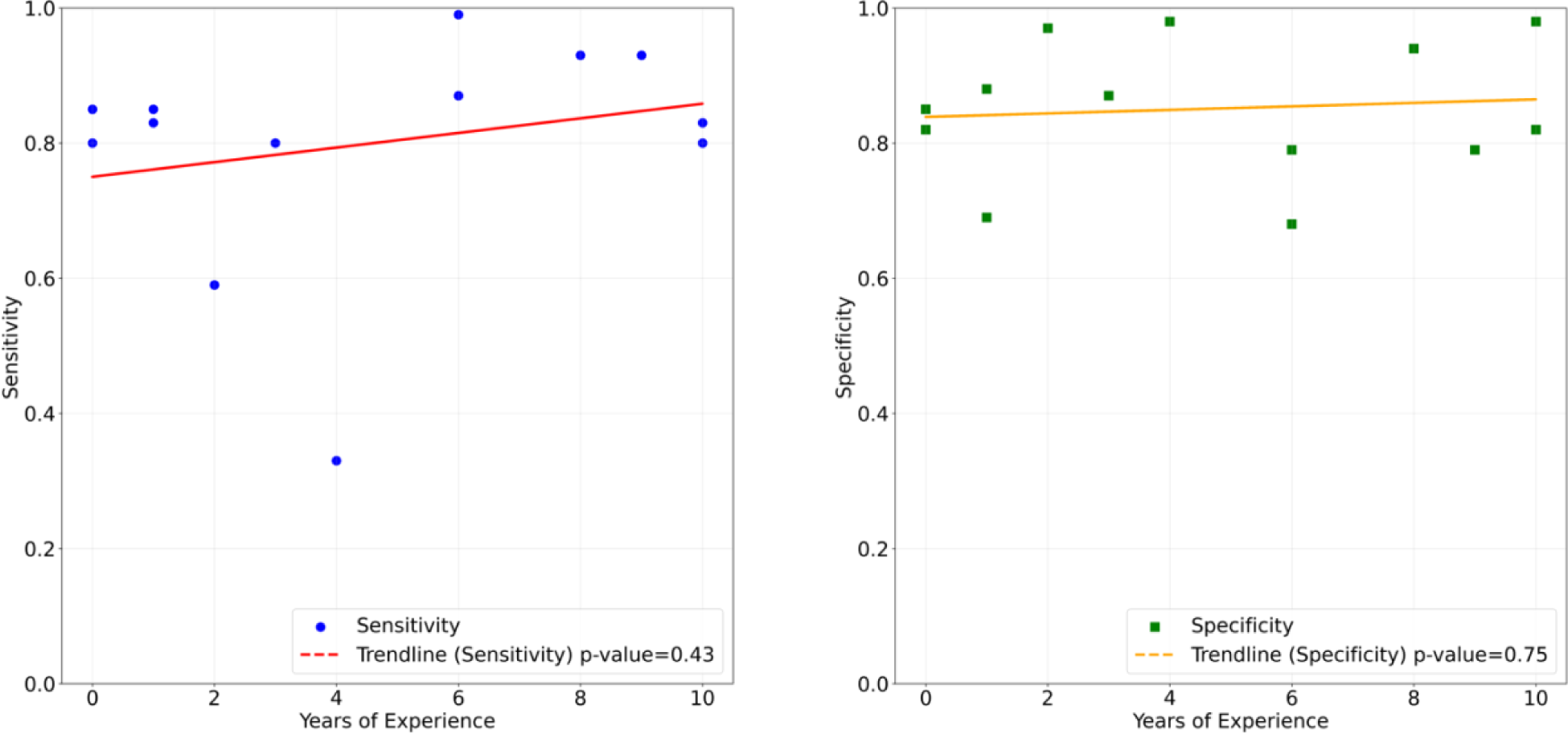
Correlation between sensitivity (left) or specificity (right) in detecting referable glaucoma and years of clinical experience among independent clinician graders.

Algorithm performance for detecting referable glaucoma on the patient level based on LAC DHS optometrist labels of the test dataset had an AUC of 0.92 (95% CI, 0.90-0.94). Individual graders, including a 4^th^ glaucoma specialist, had a sensitivity ranging from 0.32 to 0.91 and a specificity ranging from 0.61 to 0.98 (Figure 3).

**Figure 3.**
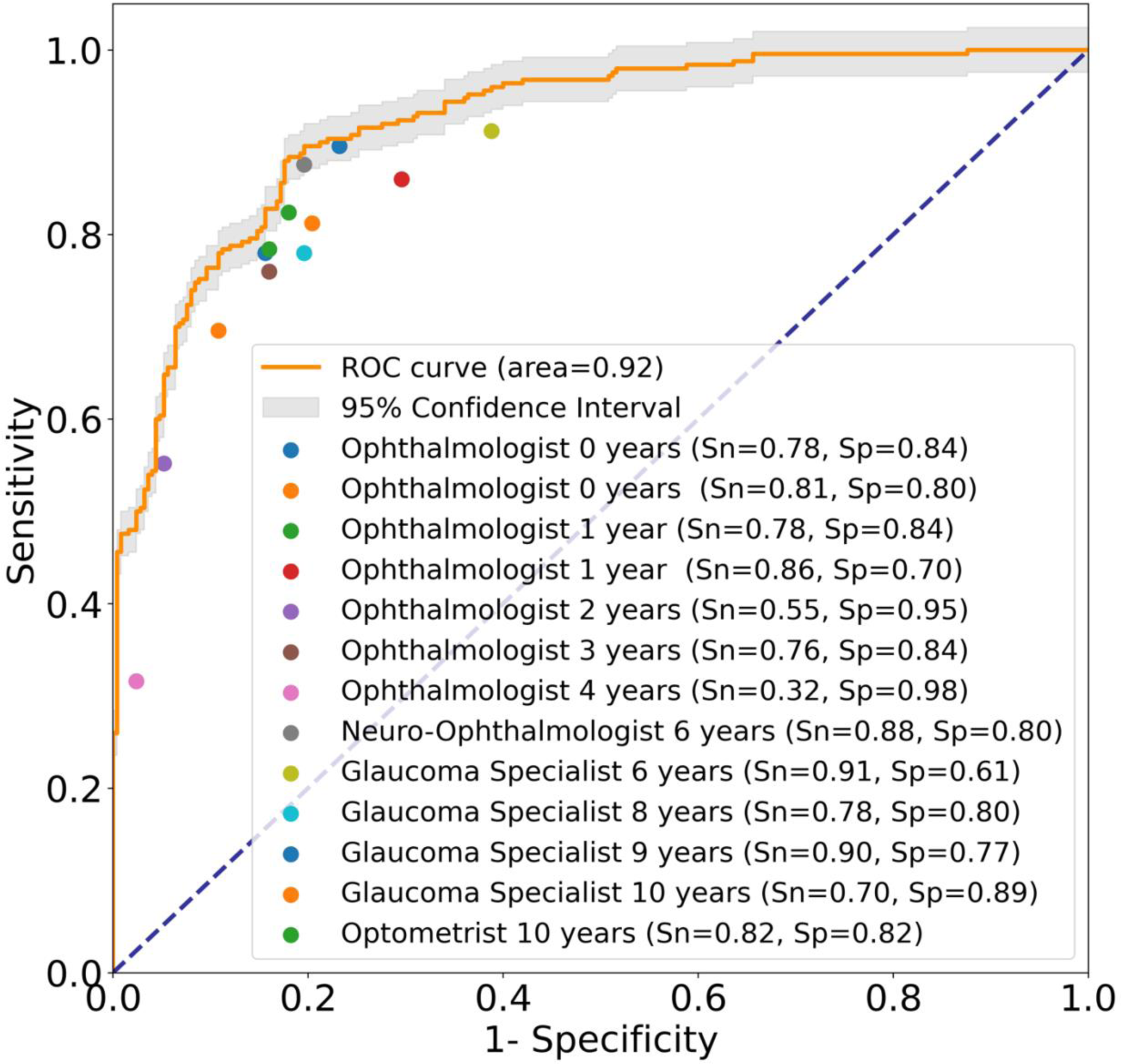
Patient-level algorithm and independent clinician performance (with years of experience) when using patient-level Los Angeles County Department of Health Services (LAC DHS) optometrist reference labels. Sn = Sensitivity; Sp = Specificity.

Algorithm performance on the eye level based on expert panel labels of the test dataset had an AUC of 0.92 (95% CI, 0.90-0.93) with a sensitivity of 0.85 and specificity of 0.83. Individual graders had a sensitivity ranging from 0.28 to 0.99 and a specificity ranging from 0.74 to 0.99, including a sensitivity of 0.90 and specificity of 0.82 by a 4^th^ glaucoma specialist (Supplementary Figure 2).

In the sub-analysis of the 6 most frequent LAC DHS optometrist graders (N = 70 to 150 images), the DL algorithm (AUC = 0.93) approximated or exceeded optometrist sensitivity (range: 0.78 to 1.0) and specificity (range: 0.32 to 0.87) in all 6 cases (Figure 4).

**Figure 4:**
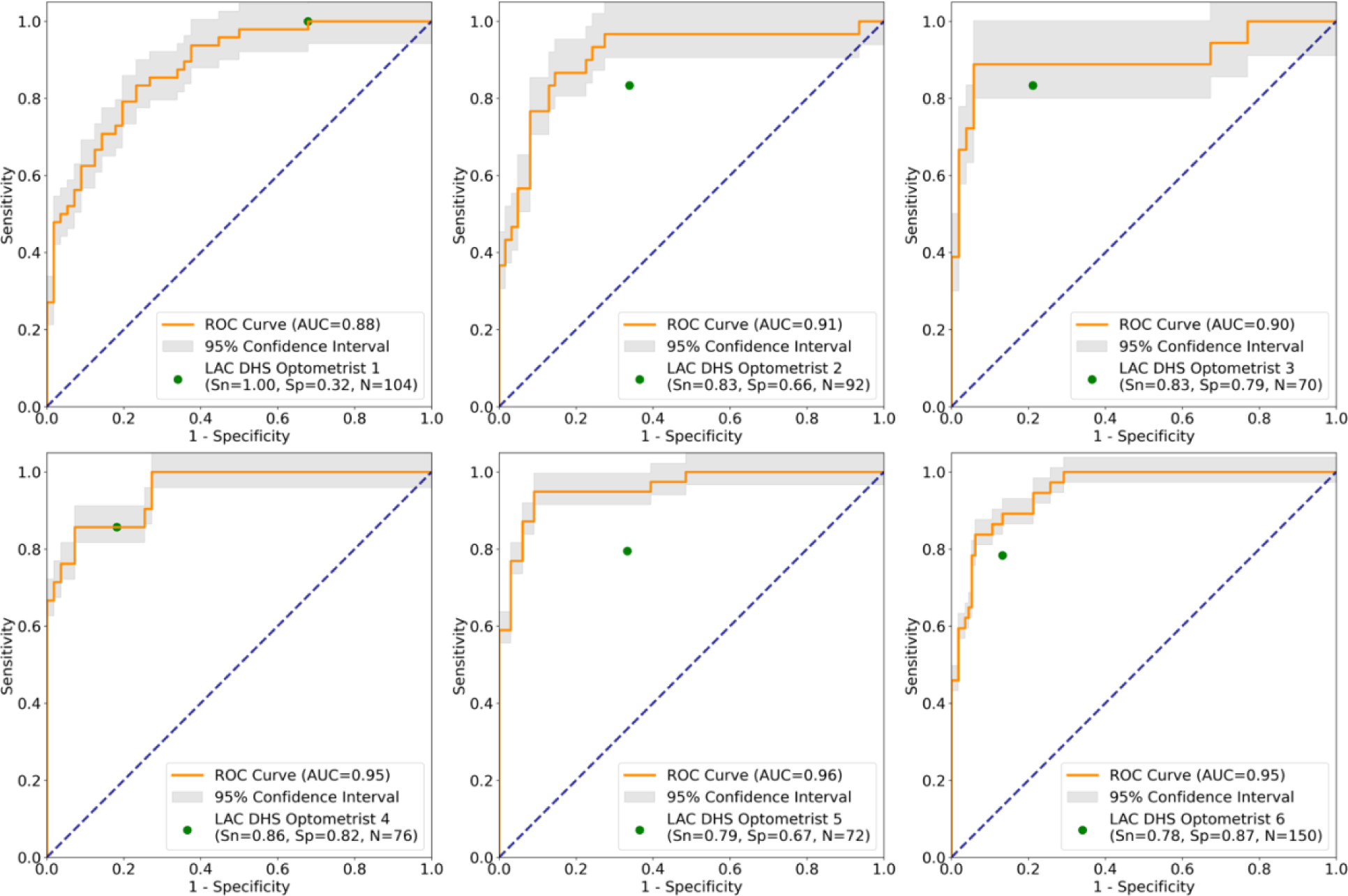
Sub-analysis of patient-level algorithm and 6 LAC DHS optometrist performance in subsets of the test dataset when using patient-level expert panel reference labels. Sn = Sensitivity; Sp = Specificity.

## Discussion

In this study, we developed a DL algorithm for detecting referable glaucoma from fundus photographs of LAC DHS teleretinal screening patients that matched or exceeded performance by clinicians with a wide range of clinical expertise, including LAC DHS optometrists and independent ophthalmologists. The algorithm, trained on patient-level labels provided by 21 trained LAC DHS optometrists, demonstrated robust performance across three sets of reference labels. In addition, LAC DHS optometrists independent ophthalmologists exhibited wide ranges of sensitivity and specificity that raise concerns about variability associated with human grading of fundus photographs. Our findings highlight potential benefits of adopting AI-based strategies to improve the reproducibility, timeliness, and scalability of glaucoma care, which could lead to earlier glaucoma detection and intervention.

While several DL algorithms for detecting referable or manifest glaucoma from fundus photographs have previously been reported, none have been as rigorously validated against standard-of-care human grading as in the current study.^10-13^ Our algorithm’s performance (AUC > 0.9) falls within the general range of performance demonstrated by these previous algorithms.^9-14^ However, it is difficult to evaluate algorithm performance based solely on comparisons with previous algorithms due to inter-study differences in disease definitions, study populations, and AI methodology. Therefore, we focused on producing a higher level of evidence to instill confidence in LAC DHS clinicians, patients, and healthcare administrators, especially given our plan to implement the algorithm in a real-world teleretinal screening environment. In a rigorous comparison with human graders, our algorithm demonstrated excellent performance, matching or exceeding the sensitivity and specificity of 13 independent clinicians with a wide range of clinical experience. In a separate sub-analysis, the algorithm also matched or outperformed 6 current LAC DHS optometrists. This robust performance compared to current standard-of-care human grading provides evidence supporting algorithm integration into existing LAC DHS teleretinal screening workflows to improve timeliness of referable glaucoma detection and re-allocate optometrist time for direct eye care.

We tested our DL algorithm using three different sets of reference labels to further assess the robustness of its performance. It is somewhat unsurprising that the algorithm matched or outperformed independent human graders when test labels were provided by the same LAC DHS optometrists who provided the training labels. However, it is interesting that the algorithm matched or outperformed independent human graders even when using test labels provided by an expert panel of three glaucoma specialists. The robust performance observed across test labels may partially stem from the diversity of training labels by 21 LAC DHS optometrists, which is likely advantageous when automating a task that is inherently variable on the individual-grader level.^26^ It may also partially stem from using reference labels provided by real-world LAC DHS optometrists rather than specially trained study graders. Using real-world training labels could help minimize the Hawthorne effect, by which individuals may modify their behaviors in response to being observed or scrutinized, thereby making the labels more applicable in real-world clinical settings.^28^ It is also interesting that our algorithm, which was trained on patient-level training labels generalized to both eyes, maintained stable performance even when tested using eye-level reference labels. This suggests that the majority of referable glaucoma was bilateral and that our approach was resistant to noisy training labels to some degree.

The high degree of variability among clinicians in referable glaucoma detection regardless of experience level presents a significant barrier for teleglaucoma screening programs. Our finding is consistent with previous studies that reported high variability among optometrists and/or ophthalmologists in grading CDR or detecting manifest glaucoma from fundus photographs.^23, 27^ This highlights an important issue associated with human grading in teleretinal screening workflows; systematic biases by graders can lead to large-scale over- or under-detection of disease, making it difficult to standardize disease detection and limiting the scalability of teleglaucoma screening overall. This variability was also not correlated with experience level, which suggests that it may be an intrinsic property of graders that is not easily modifiable, even with extensive training. In contrast to human graders, AI algorithms can be trained using collective labels provided by a large number of graders, which may help mitigate systematic biases associated with a small number of undercallers (high specificity) or overcallers (high sensitivity). AI algorithms also provide consistent and reproducible image analysis, and sensitivity and specificity can be tailored to suit the specific needs and capacities of individual healthcare systems. Therefore, the relatively unbiased, reproducible, and adaptable nature of certain AI algorithms may make them better suited for large-scale, high throughput teleretinal screening.

Our study has some limitations. First, our training data reflects the unique demographics of the communities served by LAC DHS, which may limit algorithm generalizability in other populations.^2,5^ This concern is mitigated by our primary intention to implement the algorithm locally in the LAC DHS teleretinal screening program. However, if the algorithm is implemented more widely in the future, it would likely benefit from re-tuning using data from local populations. Second, the utility of glaucoma screening in the general population remains unclear, which calls into question the role of algorithms for detecting referable glaucoma.^25^ However, LAC DHS serves a high-risk population that is predominantly Latino, which may explain why glaucoma referrals at a CDR cutoff of 0.6 are high yield; around a quarter of LAC DHS teleretinal patients detected with referable glaucoma were diagnosed with manifest glaucoma after in-office evaluation.^8^ Finally, our algorithm only evaluates single fundus photographs, which is rather simplistic compared to the comprehensive glaucoma evaluation.^8^ However, it is important to point out that we plan to implement this algorithm in resource-constrained screening environments, where the cost of expensive diagnostic tests is prohibitive and the effectiveness of fundus photography alone has been demonstrated. Nevertheless, it is important to consider future opportunities to incorporate accessible factors, such as age and race, that could improve the predictive accuracy of glaucoma referrals and minimize the burden placed on the LAC DHS health system.^16^

In conclusion, the performance of our DL algorithm for detecting referable glaucoma matched or exceeded LAC DHS optometrists and independent clinicians, including glaucoma specialists. Implementation of validated AI algorithms that approximate expert-level performance into existing clinical workflows could enhance the timeliness and quality of care while also conserving clinician time for direct patient care, which is a valuable commodity in resource-constrained healthcare systems providing care to undeserved, safety net populations.^29-31^ AI can also provide more reproducible and adaptable diagnostic capabilities, ensuring that more patients have consistent access to a higher standard of care.^21^ However, further work is needed to address technical, ethical, and legal questions surrounding AI for glaucoma care prior to wide-spread implementation.^17,18^

## Data Availability

All data produced in the present study are not available.

## Acknowledgements

This work was supported by grant R01 EY035677 and K23 EY032985 from the National Eye Institute, National Institutes of Health, Bethesda, Maryland; a DHS-USC Safety Net Innovation Award from the Southern California Clinical and Translational Science Institute; a AI4Health Award from the University of Southern California; and an unrestricted grant to the Department of Ophthalmology from Research to Prevent Blindness, New York, NY.

---

**Supplementary Figure 1:**
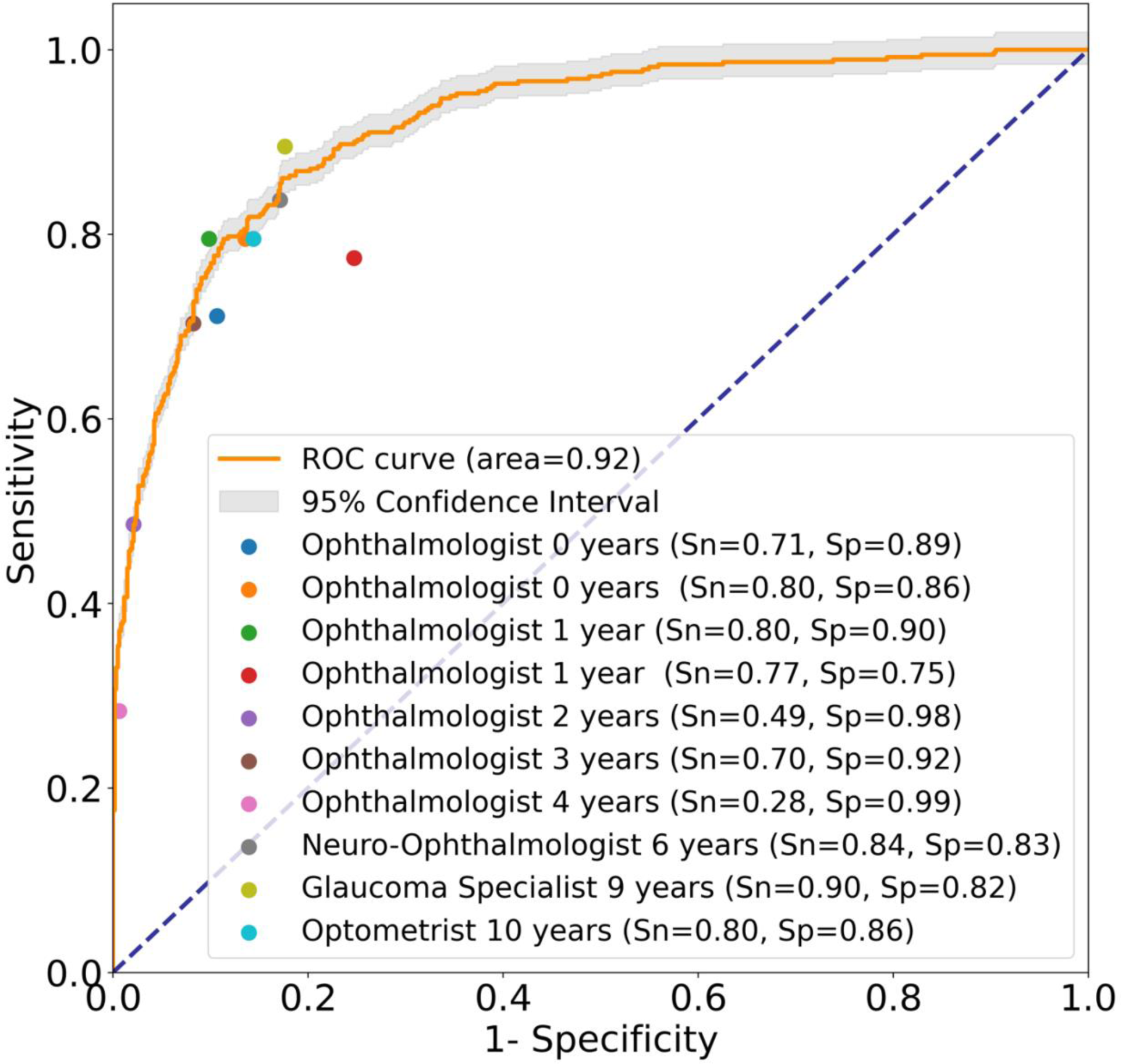
Eye-level algorithm and independent clinician performance (with years of experience) when using patient-level expert panel reference labels. Sn = Sensitivity; Sp = Specificity.

## References

1. GBD 2019 Blindness and Vision Impairment Collaborators, Vision Loss Expert Group of the Global Burden of Disease Study. Causes of blindness and vision impairment in 2020 and trends over 30 years, and prevalence of avoidable blindness in relation to VISION 2020: the Right to Sight: an analysis for the Global Burden of Disease Study. Lancet Glob Health. 2021;9(2):e144-e160. doi:10.1016/S2214-109X(20)30489-7

2. Vajaranant TS, Wu S, Torres M, Varma R. The changing face of primary open-angle glaucoma in the United States: demographic and geographic changes from 2011 to 2050. Am J Ophthalmol. 2012;154(2):303–314.e3. doi:10.1016/j.ajo.2012.02.024

3. Berkowitz ST, Finn AP, Parikh R, Kuriyan AE, Patel S. Ophthalmology Workforce Projections in the United States, 2020 to 2035. *Ophthalmology*. 2024;131(2):133-139. doi:10.1016/j.ophtha.2023.09.018

4. Tielsch JM, Sommer A, Katz J, Royall RM, Quigley HA, Javitt J. Racial variations in the prevalence of primary open-angle glaucoma. The Baltimore Eye Survey. JAMA. 1991;266(3):369–374. https://www.ncbi.nlm.nih.gov/pubmed/2056646.

5. Varma R, Wang D, Wu C, et al. Four-year incidence of open-angle glaucoma and ocular hypertension: the Los Angeles Latino Eye Study. Am J Ophthalmol. 2012;154(2):315–325.e1. doi:10.1016/j.ajo.2012.02.014

6. Barquet-Pizá V, Siegfried CJ. Understanding racial disparities of glaucoma. Curr Opin Ophthalmol. 2024;35(2):97–103. doi:10.1097/ICU.0000000000001017

7. Davuluru SS, Jess AT, Kim JSB, Yoo K, Nguyen V, Xu BY. Identifying, Understanding, and Addressing Disparities in Glaucoma Care in the United States. Transl Vis Sci Technol. 2023;12(10):18. doi:10.1167/tvst.12.10.18

8. Yuen J, Xu B, Song BJ, Daskivich LP, Rodman J, Wong BJ. Effectiveness of Glaucoma Screening and Factors Associated with Follow-up Adherence among Glaucoma Suspects in a Safety-Net Teleretinal Screening Program. Ophthalmol Glaucoma. 2023;6(3):247–254. doi:10.1016/j.ogla.2022.10.007

9. Chaurasia AK, Greatbatch CJ, Hewitt AW. Diagnostic Accuracy of Artificial Intelligence in Glaucoma Screening and Clinical Practice. J Glaucoma. 2022;31(5):285–299. doi:10.1097/IJG.0000000000002015

10. Li Z, He Y, Keel S, Meng W, Chang RT, He M. Efficacy of a Deep Learning System for Detecting Glaucomatous Optic Neuropathy Based on Color Fundus Photographs. Ophthalmology. 2018;125(8):1199–1206. doi:10.1016/j.ophtha.2018.01.023

11. Medeiros FA, Jammal AA, Mariottoni EB. Detection of Progressive Glaucomatous Optic Nerve Damage on Fundus Photographs with Deep Learning. Ophthalmology. 2021;128(3):383–392. doi:10.1016/j.ophtha.2020.07.045

12. Li F, Yan L, Wang Y, et al. Deep learning-based automated detection of glaucomatous optic neuropathy on color fundus photographs. Graefes Arch Clin Exp Ophthalmol. 2020;258(4):851–867. doi:10.1007/s00417-020-04609-8

13. Al-Aswad LA, Kapoor R, Chu CK, et al. Evaluation of a Deep Learning System For Identifying Glaucomatous Optic Neuropathy Based on Color Fundus Photographs. J Glaucoma. 2019;28(12):1029–1034. doi:10.1097/IJG.0000000000001319

14. Murtagh P, Greene G, O’Brien C. Current applications of machine learning in the screening and diagnosis of glaucoma: a systematic review and Meta-analysis. Int J Ophthalmol. 2020;13(1):149–162. doi:10.18240/ijo.2020.01.22

15. Abràmoff MD, Tarver ME, Loyo-Berrios N, et al. Considerations for addressing bias in artificial intelligence for health equity. NPJ Digit Med. 2023;6(1):170. doi:10.1038/s41746-023-00913-9

16. Thompson AC, Jammal AA, Medeiros FA. A Review of Deep Learning for Screening, Diagnosis, and Detection of Glaucoma Progression. Transl Vis Sci Technol. 2020;9(2):42. doi:10.1167/tvst.9.2.42

17. Al-Aswad LA, Ramachandran R, Schuman JS, Medeiros F, Eydelman MB, Collaborative Community for Ophthalmic Imaging Executive Committee and Glaucoma Workgroup. Artificial Intelligence for Glaucoma: Creating and Implementing Artificial Intelligence for Disease Detection and Progression. Ophthalmol Glaucoma. 2022;5(5):e16–e25. doi:10.1016/j.ogla.2022.02.010

18. Abràmoff MD, Cunningham B, Patel B, et al. Foundational Considerations for Artificial Intelligence Using Ophthalmic Images. Ophthalmology. 2022;129(2):e14–e32. doi:10.1016/j.ophtha.2021.08.023

19. Daskivich LP, Vasquez C, Martinez C Jr, Tseng CH, Mangione CM. Implementation and Evaluation of a Large-Scale Teleretinal Diabetic Retinopathy Screening Program in the Los Angeles County Department of Health Services. JAMA Intern Med. 2017;177(5):642–649. doi:10.1001/jamainternmed.2017.0204

20. Ting DSW, Cheung CY, Lim G, et al. Development and validation of a deep learning system for diabetic retinopathy and related eye diseases using retinal images from multiethnic populations with diabetes. JAMA 2017;318:2211–23.

21. Abràmoff, M.D., Lavin, P.T., Birch, M. et al. Pivotal trial of an autonomous AI-based diagnostic system for detection of diabetic retinopathy in primary care offices. npj Digital Med 1, 39 (2018). 10.1038/s41746-018-0040-6

22. Tham YC, Li X, Wong TY, Quigley HA, Aung T, Cheng CY. Global prevalence of glaucoma and projections of glaucoma burden through 2040: a systematic review and meta-analysis. Ophthalmology. 2014;121(11):2081–2090. doi:10.1016/j.ophtha.2014.05.013

23. Varma R, Steinmann WC, Scott IU. Expert agreement in evaluating the optic disc for glaucoma. Ophthalmology. 1992;99(2):215–221. doi:10.1016/s0161-6420(92)31990-6

24. Daskivich LP, Vasquez C, Martinez C, Tseng C, Mangione CM. Implementation and Evaluation of a Large-Scale Teleretinal Diabetic Retinopathy Screening Program in the Los Angeles County Department of Health Services. JAMA Intern Med. 2017;177(5):642–649. doi:10.1001/jamainternmed.2017.0204

25. Recommendation: Primary Open-Angle Glaucoma: Screening | United States Preventive Services Taskforce. May 24, 2022. Accessed July 17, 2024.

26. Gulshan V, Peng L, Coram M, et al. Development and Validation of a Deep Learning Algorithm for Detection of Diabetic Retinopathy in Retinal Fundus Photographs. JAMA. 2016;316(22):2402–2410. doi:10.1001/jama.2016.17216

27. Harper R, Reeves B, Smith G. Observer variability in optic disc assessment: implications for glaucoma shared care. Ophthalmic Physiol Opt. 2000;20(4):265–273.

28. McCambridge J, Witton J, Elbourne DR. Systematic review of the Hawthorne effect: new concepts are needed to study research participation effects. J Clin Epidemiol. 2014;67(3):267–277. doi:10.1016/j.jclinepi.2013.08.015

29. Xu BY, Chiang M, Chaudhary S, Kulkarni S, Pardeshi AA, Varma R. Deep Learning Classifiers for Automated Detection of Gonioscopic Angle Closure Based on Anterior Segment OCT Images. Am J Ophthalmol. 2019;208:273–280. doi:10.1016/j.ajo.2019.08.004

30. Xu BY, Chiang M, Pardeshi AA, Moghimi S, Varma R. Deep Neural Network for Scleral Spur Detection in Anterior Segment OCT Images: The Chinese American Eye Study. Transl Vis Sci Technol. 2020;9(2):18. Published 2020 Mar 30. doi:10.1167/tvst.9.2.18

31. Bolo K, Apolo Aroca G, Pardeshi AA, et al. Automated expert-level scleral spur detection and quantitative biometric analysis on the ANTERION anterior segment OCT system. Br J Ophthalmol. 2024;108(5):702–709. Published 2024 May 21. doi:10.1136/bjo-2022-322328

